# BCG vaccine derived peptides induce SARS-CoV-2 T cell cross-reactivity

**DOI:** 10.1101/2020.11.21.20236018

**Authors:** Peter J. Eggenhuizen, Boaz H. Ng, Janet Chang, Ashleigh L. Fell, Wey Y. Wong, Poh- Yi Gan, Stephen R. Holdsworth, Joshua D. Ooi

## Abstract

Epidemiological studies suggest that the Bacillus Calmette-Guérin (BCG) vaccine may have protective effects against coronavirus disease 2019 (COVID-19); and, there are now more than 15 ongoing clinical trials seeking to determine if BCG vaccination can prevent or reduce the severity of COVID-19 (1). However, the mechanism by which BCG vaccination can induce a severe acute respiratory syndrome coronavirus 2 (SARS-CoV-2) specific T cell response is unknown. Here, *in silico*, we identify 8 BCG derived peptides with significant sequence homology to either SARS-CoV-2 NSP3 or NSP13 derived peptides. Using an *in vitro* co-culture system, we show that human CD4+ and CD8+ T cells primed with a BCG derived peptide developed enhanced reactivity to its corresponding SARS-CoV-2 derived peptide. As expected, HLA differences between individuals meant that not all persons developed immunogenic responses to all 8 BCG derived peptides. Nevertheless, all of the 20 individuals that were primed with BCG derived peptides developed enhanced T cell reactivity to at least 7 of 8 SARS-CoV-2 derived peptides. These findings provide a mechanistic basis for the epidemiologic observation that BCG vaccination confers protection from COVID-19; and supports the use of BCG vaccination to induce cross-reactive SARS-CoV-2 specific T cell responses.

## Introduction

Severe acute respiratory syndrome coronavirus 2 (SARS-CoV-2) causes coronavirus disease 2019 (COVID-19), an infection for which no specific vaccine is currently available (2, 3). T cells are reported to be pivotal in mounting a successful immune response against COVID-19 as recovered individuals exhibit SARS-CoV-2 specific T cell memory and T cell dysfunction, and imbalance has been reported as a hallmark of severe COVID-19 (4, 5). Both CD4+ and CD8+ T cells have been implicated in COVID-19 with CD4+ T cells being broadly Th1-like by the secretion of cytokines interleukin-2 (IL-2), interferon gamma (IFN-γ) and tumour necrosis factor (TNF), and CD8+ T cells also secreting TNF and IFN-γ as well as effecting direct target cell lysis through the secretion of perforin and granzymes (6). Cross-reactive T cells between other human coronaviruses and SARS-CoV-2 have been identified, suggesting the potential role for T cell cross-protection in COVID-19 (7, 8). Here we investigated whether cross-reactive SARS-CoV-2-specific T cells can arise from Bacillus Calmette-Guérin (BCG)-derived peptide sensitization.

BCG vaccine containing live attenuated *Mycobacterium bovis*, hereafter referred to as BCG, typically vaccinates against tuberculosis (TB). It can also induce cross-protection against pathogens unrelated to TB. The cross-protective effects have shown to reduce all-cause mortality in children and respiratory tract infections in adults (9-12). One mechanism of cross-protection is through BCG epigenetically modifying innate immune cells in the form of trained innate immunity lasting up to one year (13, 14). The heterologous effect of BCG vaccination on T cells has been demonstrated in other viral infections such as murine vaccinia virus and HPV papillomatosis (15-18).

Given the heterologous effects of BCG vaccination, more than 15 clinical trials are currently underway globally to test the cross-protective effect of BCG in COVID-19, most notably the BRACE study involving 10,000 healthcare workers in Australia and the Netherlands (1). Although reports from these prospective trials are still forthcoming, large country-level epidemiological analyses have shown a negative correlation between BCG vaccination status of a country and COVID-19 disease severity or case growth (19-21).

Here we show that the observed benefits of BCG vaccination in the context of COVID-19 can be attributed, in part, to T cell cross-reactivity.

## Results and Discussion

### SARS-CoV-2 amino acid homology with BCG

T cells specific for SARS-CoV-2 are being increasingly characterised and recognised as pivotal in mounting a successful immune response to COVID-19 (6). To study the extent that BCG-primed T cells could cross-react with SARS-CoV-2 epitopes and promote viral clearance, we first performed NCBI Protein Blast searches against the SARS-CoV-2 proteome, restricting results to BCG proteins. Regions of protein sequence homology were identified between BCG sequences and the non-structural proteins NSP3 and NSP13 located in ORF1ab of SARS-CoV-2 (Fig. 1 and table S1). When processed as 15mers for MHCII presentation, these regions exhibit up to 60% identity and 73.3% similarity between BCG and SARS-CoV-2 (table S1). Percent identity and similarity of constituent 9mers for MHCI presentation are up to 88.8% and 100%, respectively, permitting cross-reactive CD4+ and CD8+ T cell responses.

**Figure 1.**
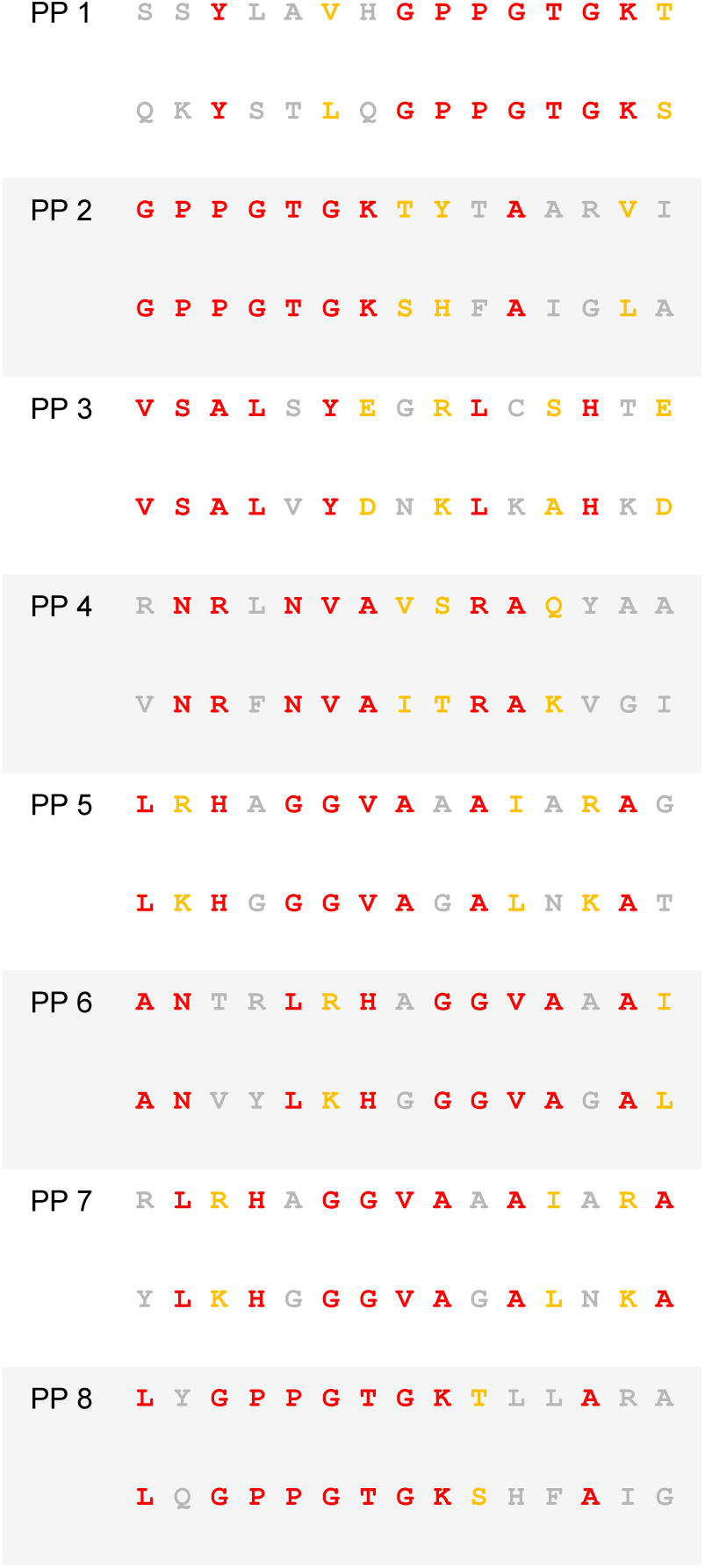
Sequence homology between BCG and SARS-CoV-2. Amino acid sequence alignment of the peptide pairs (PP) of BCG (top sequence) and SARS-CoV-2 (bottom sequence) used in this study. Red coloured amino acid – identity. Yellow coloured amino acid – similarity. Grey coloured amino acid – no identity or similarity.

NSP3 is a papain-like proteinase that shares a macro domain with the BCG proteins: macro-domain-containing protein and UPF0189 protein. This macro-domain-containing protein is conserved among the *Mycobacterium tuberculosis* complex including BCG (accession number WP_003909539.1). NSP13 is a helicase that shares homology with BCG proteins RecB nuclease and zinc-metalloprotease-FtsH. Both RecB nuclease and zinc-metalloprotease-FtsH contain a walker-A-motif sequence that is identical in NSP13 of SARS-CoV-2. Additionally, RecB nuclease contains two other regions of homology with NSP13 around amino acid residues 952-966 and 1093-1107. As has been previously reported, NSP13 is highly conserved between other human coronaviruses. Thus, the T cell cross-protective potential of BCG holds not only for SARS-CoV-2 as we have shown but potentially with other human coronaviruses that cause the common cold (229E, NL63, OC43 and HKU1) and the more serious human coronaviruses SARS-CoV and Middle East respiratory syndrome coronavirus (MERS-CoV). NSP3 is, however, not as widely conserved among coronaviruses (7, 22).

In order for cross-reactivity to occur between T cells that share epitope homology, a significant degree of homology must also be paired with the capacity of an immunogenic peptide to bind cognate MHC class I or II. Indeed, HLA binding has been reported as important in COVID-19 severity. Patients with mild COVID-19 presented MHCI molecules with a higher theoretical affinity than those with moderate to severe COVID-19 (23). To assess the capacity of BCG epitopes to bind HLA alleles that broadly cover the global population, we performed *in silico* prediction analyses of peptide-MHC binding affinity using NetMHCIIpan 4.0 and NetMHCpan 4.1 across each region of homology as 9mers or 15mers overlapping by 1 amino acid residue in MHCI and MHCII binding, respectively (24). HLA alleles in the analysis were selected based on previously reported reference sets giving maximal global population coverage (25, 26). We found that the BCG derived peptides with homologous sequences to SARS-CoV-2 peptides exhibited broad MHC class II and MHC I binding capacity (Figs S1 and S2).

To determine the cross-reactive immunogenicity of these BCG derived peptides across diverse HLA-types, we selected 10 healthy HLA-typed blood donors with different HLA types (Table S2b). Based on IEDB population coverage, our collection of HLA-typed individuals gave a global MHC Class I and II coverage of 97.21% and 99.97%, respectively (27). In addition, binding affinity predictions of the homologous peptides to HLA alleles from the 10 HLA-typed donors used in this study were analysed (Figs. S1 & S2). Based on homology and strong binding, a selection of eight different 15mer peptide pairs (PP1-8) were chosen for subsequent experimentation on human donors (Fig. 1). To determine if HLA-typing was necessary, we also tested the cross-reactive immunogenicity of the BCG derived peptides on 10 non-HLA-typed persons.

### T cell cross reactivity

To determine if priming with BCG peptide enhances T cell responses to SARS-CoV-2 peptides, we compared CD4+ and CD8+ T cell responses to SARS-CoV2 peptides using cells that were either primed with a control peptide (invariant chain peptide, CLIP) or BCG peptide. CD3+ T cells were isolated from donors (n=20, table S2a) and co-cultured with dendritic cells (DCs) *in vitro* (Fig. 3). Individual BCG peptides were first used to sensitize and expand the BCG-specific T cells, simulating a BCG vaccination. T cells were then rested for two days without antigen stimulation then restimulated with the SARS-CoV-2 peptides. To measure T cell responses, we performed intracellular cytokine staining (ICS) for IFN-γ, TNF, IL-2, perforin; surface staining for the early T cell activation marker CD69, and a two-colour proliferation assay to differentiate between a primary proliferative and secondary proliferative response using a combination of both Cell Trace Yellow (CTY) and Cell Trace Violet (CTV). A positive response was defined as an increase compared to control.

**Figure 2.**
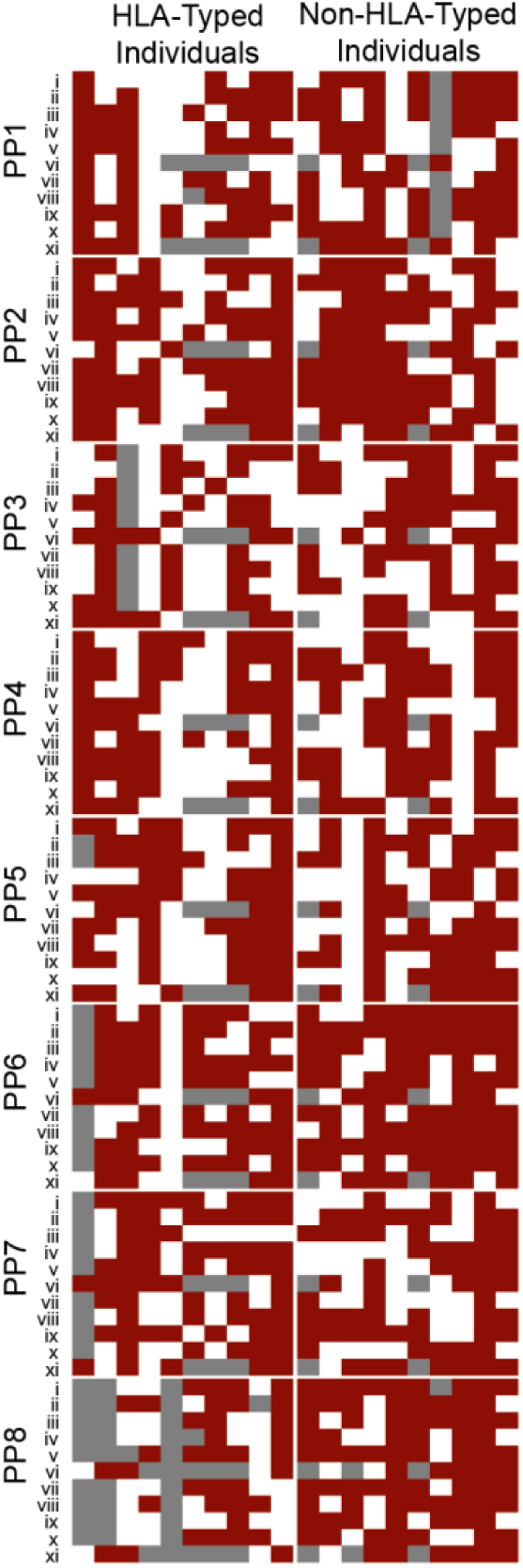
BCG induces broad cross-reactive T cell responses across individuals. Heat map of individuals representing global HLA coverage shows improved SARS-CoV-2 T cell responses when stimulated with SARS-CoV-2 peptide. Individual donor T cell responses to the 8 peptide pairs (PP1-PP8) across 11 parameters (i-xi) determined by flow cytometry. i – CD8+ IFN-γ, ii – CD8+ TNF, iii – CD8+ IL-2, iv – CD8+ CD69, v – CD8+ Perforin, vi – CD8+ proliferation, vii – CD4+ IFN-γ, viii – CD4+ TNF, ix – CD4+ IL-2, x – CD4+ CD69, xi – CD4+ proliferation. A responder (red) is defined as showing a positive response after subtraction of the control primed response to SARS-CoV-2. A non-responder in white is defined as showing no positive staining after subtraction of the control response. Grey – data not available. Individuals were grouped by known or unknown HLA-type highlighting similar patterns between the two groups.

**Figure 3.**
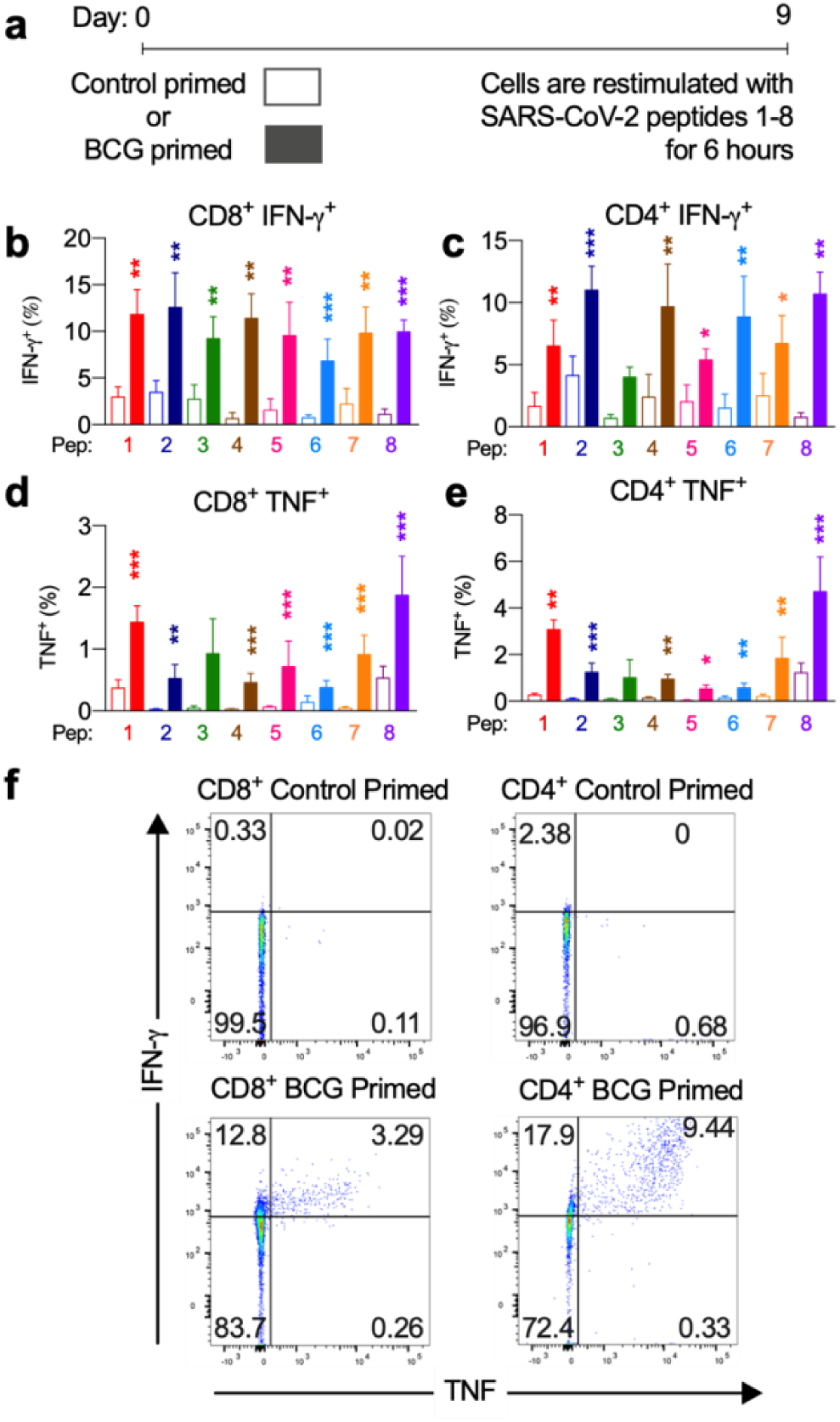
BCG priming enhances CD4+ T cell and CD8+ T cell responses against SARS-CoV-2. BCG-peptide primed CD3+ T cells were restimulated with SARS-CoV-2-peptide-pulsed dendritic cells for 6 hr and analysed by intracellular cytokine staining. Unshaded bars - Control primed using irrelevant peptide CLIP_103-117_ then SARS-CoV-2-peptide 1-8 restimulated. Shaded bars – BCG peptide 1-8 primed then SARS-CoV-2-peptide homologue restimulated. a) Brief timeline of the culture, b) CD8+ IFN-γ+ responses (*n*=9-12), c) CD4+ IFN-γ+ responses (*n*=5-13), d) CD8+ TNF+ responses (*n*=4-14), e) CD4+ TNF+ responses (*n*=6-16), f) Representative TNF (x-axis) and IFN-γ (y-axis) dot plots of a responder donor with their corresponding SARS-CoV-2 primary response control. \**P* < 0.05, \*\**P* < 0.01, \*\*\**P* < 0.001 by Wilcoxon matched-pairs signed rank test of responder samples, comparing the magnitude of response to SARS-CoV-2 peptides with or without BCG peptide priming.

All individuals (n=20) exhibited a positive response to at least 7 out of 8 SARS-CoV-2 peptides (Fig. 2). The enhanced positive cross-reactive response confirms the prediction of high HLA binding affinity and we confirm these cross-reactive peptides are immunogenic as they elicit CD4+ Th1-like responses and robust CD8+ responses.

Next, we assessed the degree of SARS-CoV-2 T cell reactivity enhancement conferred by BCG priming compared to control primed T cells (Fig. 3 and Figs. S5 & S6). In CD8+ cytotoxic T cells, IFN-γ, TNF and IL-2 cytokine production across all 8 peptide pairs significantly increased (Fig. 3 and Fig. S5a). The IFN-γ mean fold increase in expression ranged from 2.3-fold (PP3) to 16.3-fold (PP4). Mean fold increase in TNF expression from CD8+ cells ranged from 1.7-fold (PP6) to 23.9-fold (PP2). IL-2 production from CD8+ cells showed a mean fold increase from 3.1-fold (PP2) to 33.1-fold (PP5).

CD4+ T helper cells exhibited similarly significantly increased IFN-γ, TNF and IL-2 production across all 8 peptide pairs (Fig. 3 and Fig. S6a). In CD4+ cells from responder individuals, IFN-γ mean fold increase in expression ranged from 1.7-fold (PP2, PP5 & PP7) to 12.9-fold (PP8). Mean fold increase in TNF from CD4+ cells ranged from 2.9-fold (PP8) to 14.1-fold (PP2 & PP5). IL-2 production from CD4+ cells showed a mean fold increase from 2.1-fold (PP5) to 12.3-fold (PP8).

Patterns of cytokine production were varied between individuals and peptide pairs which is reflected in the complex pattern of T cell cytokine expression and phenotypes that BCG vaccination is known to produce (28). Indeed, the IFN-γ and TNF response was mixed with some individuals making only IFN-γ or TNF in response to a particular peptide pair and some being positive for both (Fig. S8). This observation is concordant with previously reported responses in COVID-19 (6, 7).

Since the COVID-19 CD8+ response involves the secretion of perforin and granzymes for an effective antiviral response, we measured perforin expression by ICS. We found that CD8+ T cells primed with BCG-derived peptides had an enhanced perforin expression upon SARS-CoV-2 restimulation when compared to control primed cells (Fig. S5b). Cross-reactive perforin expression in responders was significantly increased across all 8 peptide pairs with a mean fold-increase ranging from 1.9-fold (PP1) to 47.2-fold (PP4). Thus, cross-reactive CD8+ T cells can effect an antiviral response by target cell lysis.

In order to mount an effective T cell response to COVID-19, antigen-specific T cells need to become activated and undergo clonal expansion. To assess whether T cells pre-stimulated with BCG-derived peptides exhibit enhanced T cell activation when restimulated with SARS-CoV-2 homologues, expression of early T cell activation marker CD69 was assessed by flow cytometry. We show that when compared to a SARS-CoV-2 primary response, the BCG primed T cells increased CD69 expression across all 8 peptide pairs (Figs. 5c, 6c). CD69 expression in responders showed a mean fold-increase ranging from 3.2-fold (PP1) to 29.6-fold (PP5) for CD4+ cells and from 1.7-fold (PP1) to 10.5-fold (PP2) for CD8+ cells. These cross-reactive T cells that show increased activation when primed with BCG peptides and restimulated with SARS-CoV-2 homologues, are able to proliferate and produce superior effector functions than those not presensitized with BCG peptides. This may be of importance in swift and effective viral clearance of SARS-CoV-2 in COVID-19 patients.

To assess whether T cells primed with BCG derived peptides show enhanced T cell proliferation upon SARS-CoV-2 peptide restimulation, cell proliferation dye CTY was used to assess the proliferation after BCG priming followed by CTV to assess the proliferation after SARS-CoV-2 restimulation. All donor samples primed with a BCG peptide developed enhanced T cell proliferation to at least 3 out of the 8 SARS-CoV-2 peptides tested (Fig. 2). The magnitude of the enhanced proliferative response was also assessed in BCG-primed individuals who responded SARS-CoV-2 restimulation. Specifically, we compared the SARS-CoV-2 peptide induced proliferation in cells that were first sensitized with BCG peptide or with control peptide. In all of the tested peptide pairs (PP1-PP8) and across both CD4+ and CD8+ T cells, BCG peptide sensitized cells developed significantly enhanced proliferation to its SARS-CoV-2 homologous peptide (Fig. 4). In the responders, T cell proliferation was enhanced in CD8+ T cells between 19% (PP3 and PP5) to 51% (PP6) and in CD4+ T cells by 11% (PP5) to 39% (PP8). Therefore, we show that BCG peptides have the ability to cross-protect against SARS-CoV-2 by T cell activation and heightened T cell proliferation.

**Figure 4.**
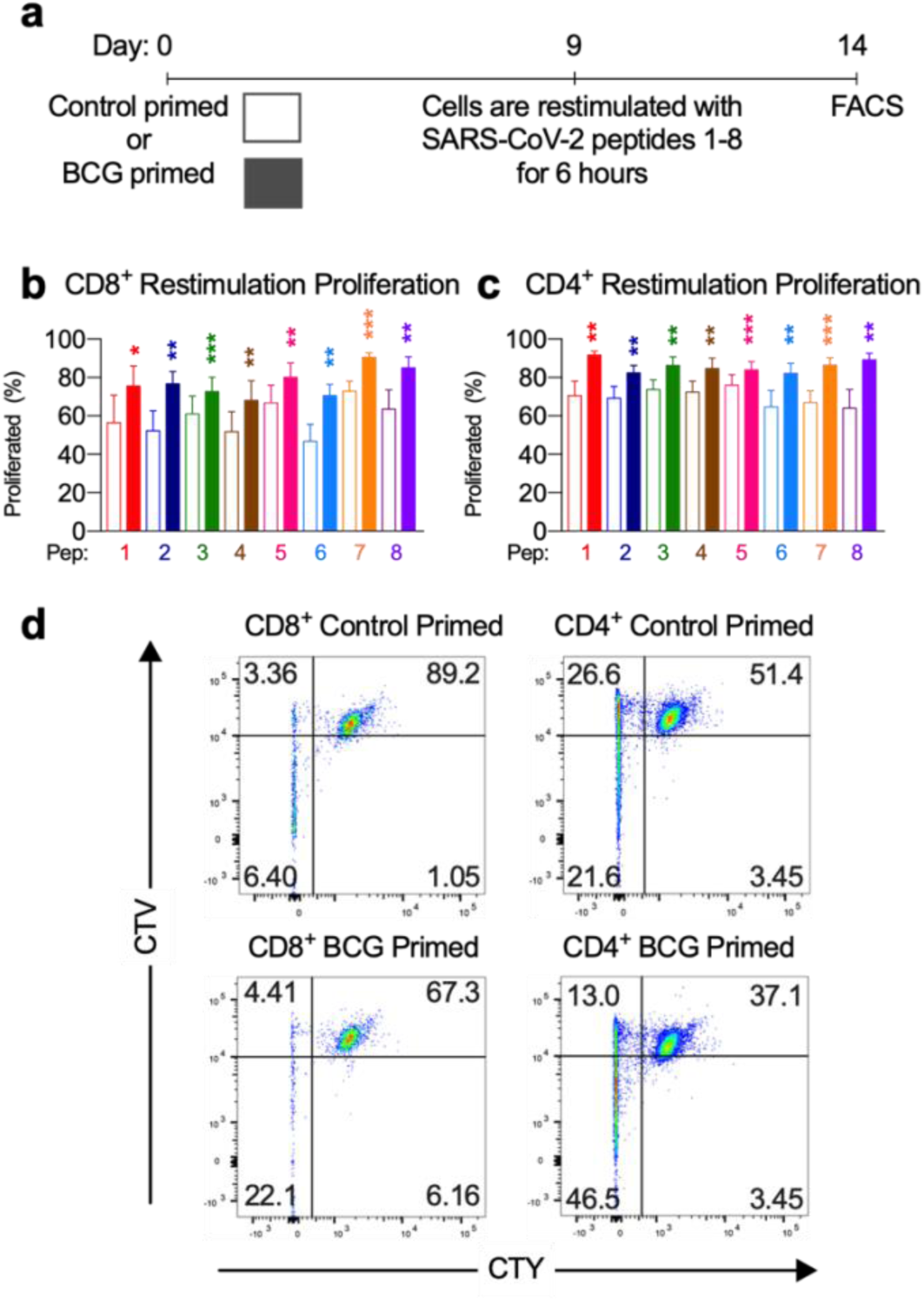
BCG priming enhances CD4+ and CD8+ T cell proliferation. a) Brief culture timeline. b) CD8+ restimulation proliferation response is enhanced by BCG priming (*n*=9-12). c) CD4+ restimulation proliferation response is enhanced by BCG priming (*n*=6-11). Unshaded bars – Control primed using irrelevant peptide CLIP_103-117_ then SARS-CoV-2-peptide restimulated. Shaded bars – BCG peptide primed then SARS-CoV-2-peptide homologue restimulated. d) Representative CTV versus CTY dot plots of CD4+ and CD8+ cultured cells indicating proliferation. Top right quadrant gate of CTY^hi^CTV^hi^ cells did not proliferate upon priming or restimulation. Top left quadrant gate of CTY^lo^CTV^hi^ cells did proliferate upon priming but not with restimulation. Bottom left quadrant gate of CTY^lo^CTV^hi^ cells proliferated upon both priming and restimulation. \**P* < 0.05, \*\**P* < 0.01, \*\*\**P* < 0.001 by Wilcoxon matched-pairs signed rank test of responder samples, comparing the magnitude of response to SARS-CoV-2 peptides with or without BCG peptide priming.

To confirm and establish the T memory cell phenotype of proliferated T cells after BCG stimulation, the proportion of T effector memory (Tem), T central memory (Tcm) and T effector memory re-expressing CD45RA (TEMRA) cells were determined based on CD45RA and CCR7 expression patterns on the proliferated CD4+ or CD8+ cells (Fig. S7). Across all 8 tested BCG peptides, greater than 99% of proliferated BCG-stimulated cells exhibited Tcm, Tem or TEMRA memory phenotype at day 16, the rest being naïve phenotype (CD45RA+, CCR7+). Of these memory cells, Tem was the predominant phenotype of CD4+ and CD8+ cells. CD4+ cells exhibited a minor subpopulation of Tcm and few TEMRA and CD8+ cells exhibited a minor subpopulation of TEMRA and few Tcm. Therefore, T memory phenotypes predominate in BCG-stimulated T cells providing an explanation for their potential to heighten recall responses.

Although the self-reported BCG vaccination status of our donors was known (*n*=10), we found no significant difference in responses from BCG vaccinated individuals compared to unvaccinated individuals (data not shown). The co-culture assay was not designed to test the direct *ex vivo* recall response of prior BCG vaccination but rather to simulate vaccination *in vitro* by pre-stimulating with BCG-derived peptides. We analysed an equal number of males and females in this study (*n*=10 each) and no significant sex-specific differences were found in the parameters measured (data not shown).

## Conclusions

Collectively, our results demonstrate that CD4+ and CD8+ T cells specific for BCG derived peptides are cross-reactive with SARS-CoV-2 peptides. These data provide a mechanistic explanation for the observed negative epidemiological associations between BCG vaccinations and COVID-19 severity and mortality and support the continuation of clinical trials around the world, particularly in people at high-risk of contracting SARS-CoV-2.

## Materials and Methods

### Ethics statement

The study was conducted according to the Declaration of Helsinki and approved by Monash University Human Research Ethics Committee project ID 25834. All donors provided written informed consent.

### Data reporting

No statistical methods were used to pre-determine sample size. The experiments were not randomised. The investigators were not blinded to allocation during experiments and assessment of outcomes.

### HLA typing

Seven donors underwent high resolution class I and II molecular sequence-based typing performed by the Australian Red Cross Victorian Transplantation and Immunogenetics Service by next-generation sequencing. Three donors underwent low-resolution HLA-DR typing at the same provider. HLA typing results are contained within table S2b.

### Human sample collection

All donors provided written informed consent. Donors with prior known TB or COVID-19 infection were excluded. Donors with current or recent symptoms of COVID-19 or who tested positive for serum IgG or IgM SARS-CoV-2 antibodies by SARS-CoV-2 Colloidal Gold Immunochromatography Assay Kit (MyBioSource) were excluded. Prior BCG vaccination status was self-reported. Twenty donors were recruited into the study and their details summarized in table S2a,b. Whole blood was collected by venepuncture into K2EDTA vacutainer tubes (BD) for cell isolation or serum separator tubes (BD) for autologous serum collection.

### HLA binding prediction and allele coverage

Global allele coverage of HLA-typed donors was assessed at IEDB Analysis Resource – Population Coverage (27). NetMHCpan-4.1 and NetMHCIIpan-4.0 were used to predict binding affinity of homologous peptides to a globally representative collection of MHCI or MHCII alleles plus the alleles of our HLA-typed donors using artificial neural networks (24-26). For each region of homology, 9mers for MHCI and 15mers for MHCII overlapping by 1 amino acid underwent affinity analysis. Affinity rank was generated that normalizes prediction score by comparing to prediction of a set of random peptides. An affinity rank score of < 2 was called a strong binder. An affinity rank score of ≥ 2 and 10 was called a binder. An affinity rank score of > 10 was called a non-binder.

### Sequence alignment

Protein BLAST search of the SARS-CoV-2 proteome (sequence ID NC_045512.2) restricted to *Mycobacterium bovis* (BCG) was performed using the NCBI blastp suite (https://blast.ncbi.nlm.nih.gov/Blast.cgi). Protein sequences from SARS-CoV-2 NSP3 (YP_009725299.1), SARS CoV-2 NSP13 (YP_009725308.1), BCG RecB nuclease (KAF3412556.1), BCG UPF0189 protein (AHM07651.1), BCG Macro domain containing protein (WP_003909539.1), BCG zinc metalloprotease FtsH (AMC52863.1) and human CLIP (NP_001020330.1) were obtained from the NCBI Database (https://www.ncbi.nlm.nih.gov/protein/). Sequence alignment of SARS-CoV-2 and BCG homologues was performed using EMBOSS Needle Pairwise Sequence Alignment (29).

### Peptides

15mer peptides were synthesised with an N-terminal free amine (H-) and a free acid group at the C-terminus (-OH). Peptides were ≥ 90% pure as assessed by reversed-phase high-performance liquid chromatography (RP-HPLC) (Mimotopes). Peptide sequences used in this study can be found in table S1 and control peptide CLIP_103-117_ (PVSKMRMATPLLMQA). Lyophilized peptide was reconstituted in sterile MilliQ water with 5% (v/v) DMSO (Sigma). Final concentration of peptides used in culture was 10μg/mL and final concentration of DMSO in the cultures was 0.005% (v/v).

### Monocyte derived DC production

Human PBMCs were freshly isolated from whole donor blood in K2EDTA anticoagulant Vacutainers (BD) using Lymphoprep density gradient medium (Stemcell) and SepMate tubes (Stemcell). PBMCs were enumerated in a haemocytometer with trypan blue 0.4% (Sigma) and the CD14+ CD16-monocytes were then magnetically separated using EasySep Human Monocyte Isolation Kit and EasySep Magnet following manufacturer’s instructions (Stemcell). Freshly isolated monocytes were then enumerated in a haemocytometer with 0.4% trypan blue and differentiation culture was established to differentiate the monocytes into dendritic cells using ImmunoCult Dendritic Cell Culture Kit following instructions of the manufacturer (Stemcell). According to the protocol (Stemcell), immature DCs used in the ICS co-culture did not receive maturation supplement on day 5 of culture and mature DCs used in the proliferation and memory co-culture received maturation supplement on day 5 of culture. After 7 days culture, immature DCs were used for the ICS co-culture and mature DCs were used for the proliferation co-culture.

### T cell isolation

Human CD3+ T cells were isolated from fresh whole donor blood in K2EDTA tubes using RosetteSep HLA T Cell Enrichment Cocktail according to instructions of the manufacturer (Stemcell). Isolated CD3+ T cells were enumerated in a haemocytometer with 0.4% trypan blue (Sigma). CD3+ T cells were then used in the ICS and proliferation co-cultures.

### ICS co-culture setup

ICS co-culture was initiated with 100,000 freshly isolated human CD3+ T cells, 10,000 human immature monocyte-derived DCs and 10ug/mL of BCG peptide from PP1-8 (Fig. 1) or control peptide CLIP_103-117_ in a 96 well round-bottom plate (Corning) at 100uL per well of complete RPMI (Gibco) supplemented with 10% autologous human serum, 100 U/mL penicillin and 0.1 mg/mL streptomycin (Gibco), 2mM L-glutamine (Gibco) and 50μM 2-mercaptoethanol (Sigma). Positive assay control received anti-human CD2, anti-human CD3, and anti-human CD28 coated MACS iBeads at a ratio of 1 bead:2cells prepared from the human T cell activation/expansion kit as per the manufacturer’s instructions (Miltenyi). Negative assay control received no peptides. Co-culture was incubated at 37°C in a CO_2_ incubator (Binder). Five days later, the co-culture was supplemented with 40IU/mL recombinant human IL-2 (Stemcell) and reincubated. On day 7 of co-culture, cells were rested by washing twice in 250uL PBS to remove peptides and resuspended in 100uL fresh complete RPMI formulated as above with no peptides and reincubated. On day 9 of co-culture, cells were restimulated by washing twice with 250μL PBS then 10,000 freshly-cultured, immature DCs were added per well with 10μg/mL of SARS-CoV-2 peptide from PP1-8 (Fig. 1) or control peptide CLIP_103-117_ and 1μg/mL anti-human CD28 monoclonal antibody (clone CD28.2, eBioscience) in serum-free RPMI. Positive assay control received anti-human CD2, anti-human CD3, and anti-human CD28 coated MACS iBeads at a ratio of 1 bead:2cells. Negative assay control received no peptides. To pulse the DCs with peptide, co-culture was incubated for 2 hours at 37°C in a CO_2_ incubator (Binder). After 2 hours, media was adjusted to contain 10% autologous serum and 1X protein transport inhibitor cocktail containing brefeldin A and monensin (eBioscience) was added and reincubated. After 6 hours at 37°C in a CO_2_ incubator, cells were harvested for flow cytometric analysis by ICS. The entire culture system was setup to be autologous.

### Proliferation co-culture setup

Proliferation co-culture was initiated with 100,000 freshly isolated CD3+ T cells stained with cell proliferation dye Cell Trace Yellow according to the manufacturer (Invitrogen), 10,000 human mature DCs and 10μg/mL of BCG peptide from PP1-8 (Fig. 1) or control peptide CLIP_103-117_ in a 96 well round-bottom plate (Corning) at 100uL per well of complete RPMI (Gibco) supplemented with 10% autologous human serum, 100 U/mL penicillin and 0.1 mg/mL streptomycin (Gibco), 2mM L-glutamine (Gibco) and 50μM 2-mercaptoethanol (Sigma). Positive assay control received anti-human CD2, anti-human CD3, and anti-human CD28 coated MACS iBeads at a ratio of 1 bead:2cells prepared from the human T cell activation/expansion kit as per the manufacturer’s instructions (Miltenyi). Negative assay control received no peptides. Co-culture was incubated at 37°C in a CO_2_ incubator (Binder). Seven days later, cells were washed twice in 250uL PBS to remove peptides and resuspended in 100uL complete RPMI formulated as above with no peptides and reincubated. On day 9 of co-culture, cells were washed twice in 250uL PBS and stained with Cell Trace Violet cell proliferation dye according to the manufacturer’s instructions (Invitrogen). Then 10,000 freshly-cultured, human mature DCs were added per well with 10μg/mL of SARS-CoV-2 peptide from PP1-8 (Fig. 1) or control peptide CLIP_103-117_. Positive assay control received anti-human CD2, anti-human CD3, and anti-human CD28 coated MACS iBeads at a ratio of 1 bead:2cells. Negative assay control received no peptides. Co-culture was incubated at 37°C in a CO_2_ incubator for 7 days then harvested for flow cytometric analysis. The entire culture system was setup to be autologous.

### ICS flow cytometry staining and analysis

After culturing, cells were stained with Live/Dead Fixable Near Infra-Red Dead Cell Stain Kit according to the manufacturer’s instructions (Invitrogen). Cells were then stained with surface markers anti-human CD3 Brilliant Violet 510 (clone OKT3, Biolegend), anti-human CD4 APC (clone OKT4, eBioscience), anti-human CD8 Alexa Fluor 488 (clone HIT8a, Biolegend) and anti-human CD69 Brilliant UV 395 (clone FN50, BD). After surface staining, cells were fixed and permeabilized with Transcription Factor Staining Buffer Set according to the manufacturer’s instructions (eBioscience). Cells were subsequently stained for intracellular markers with anti-human IFN-γ PE Cy7 (clone 4S.B3, eBioscience), anti-human TNF Brilliant Violet 421 (clone Mab11, Biolegend), anti-human IL-2 Brilliant Blue 700 (clone MQ1-17H12, BD) and anti-human perforin PE (clone B-D48, Biolegend). Single colour controls were prepared using UltraComp eBeads (Invitrogen) for single colour control antibodies and ArC amine reactive compensation bead kit (Invitrogen) for Live/Dead single colour control. After staining, cells were resuspended in PBS and acquired on an LSR-Fortessa X20 flow cytometer (BD) using BD FACSDiva software version 8.0.1. Samples were analyzed in FlowJo 10.6.2. FMO controls were used to determine positive gating (Fig. S3).

Individuals that responded in the given parameters to SARS-CoV-2 after BCG priming (Fig. 2) were defined as showing positive staining after subtraction of the primary SARS-CoV-2 response control (CLIP_103-117_ primed, SARS-CoV-2 peptide stimulated). A non-responder was defined as showing no positive staining after subtraction of the primary SARS-CoV-2 response control. For statistical analysis (Fig. 3, Figs. S5 & S6), the responders were selected as those with positive staining after subtraction of the primary SARS-CoV-2 response control or restimulation background control (BCG primed, irrelevant peptide CLIP restimulated). The responders then had the restimulation background control (BCG primed and CLIP_103-117_ restimulated) subtracted from the corresponding BCG primed, SARS-CoV-2 test sample and the primary SARS-CoV-2 response control to remove any assay related background stimulation.

### Proliferation flow cytometry staining and analysis

After culturing, cells were stained with Live/Dead Fixable Near Infra-Red Dead Cell Stain Kit according to the manufacturer’s instructions. Cells were then stained with surface markers anti-human CD3 PerCP (clone SK7, Biolegend), anti-human CD4 APC (clone OKT4, eBioscience), anti-human CD8 Alexa Fluor 488 (clone HIT8a, Biolegend), anti-human CD45RA Brilliant Violet 711 (clone HI100, Biolegend) and anti-human CCR7 Brilliant UV (clone 3D12, BD). Single colour controls were prepared using UltraComp eBeads (Invitrogen) for single colour control antibodies, ArC amine reactive compensation bead kit (Invitrogen) for Live/Dead single colour control and Cell Trace Violet and Cell Trace Yellow single stained co-cultured cells along with unstained co-cultured cells. After staining, cells were resuspended in 0.5% BSA, 2mM EDTA/PBS and acquired on an LSR-Fortessa X20 flow cytometer (BD) using BD FACSDiva software version 8.0.1. .fcs files were analysed in FlowJo 10.6.2. All fluorescence based gating except CTY and CTV is determined based on fluorescence minus one (FMO) controls (Fig. S4). CTV and CTY gating is based on the point at which the first cell division took place visible by fluorescence dye dilution. Individuals that showed a proliferation response when BCG primed, SARS-CoV-2 restimulated (Fig. 2) were defined as showing positive staining after subtraction of the primary SARS-CoV-2 response control (CLIP_103-117_ primed, SARS-CoV-2 peptide stimulated). A non-responder was defined as showing no positive staining after subtraction of the primary SARS-CoV-2 response control. For statistical analysis (Fig. 4), the responders were selected as those with positive staining after subtraction of the primary SARS-CoV-2 response control or restimulation background control (BCG primed, irrelevant peptide CLIP restimulated). Proliferation in response to BCG priming and SARS-CoV-2 restimulation was calculated as the proportion of CD4+ or CD8+ cells that underwent proliferation post-priming and post-restimulation (CTY^lo^CTV^lo^) of total proliferated cells (CTY^lo^CTV^lo^ and CTY^lo^ CTV^high^).

### Statistics

Flow cytometry data was exported from FlowJo 10.6.2 (BD) and analysed using R Studio version 1.3.959 before being analysed in GraphPad Prism 7 (Graphpad Software Inc.). A Shapiro Wilk test was used to determine normality followed by a two-tailed, Wilcoxon matched-pairs signed rank test to compare the responses of BCG primed with control primed samples from responders.

## Data Availability

Correspondence and requests for materials should be addressed to Dr. Ooi.

## Author Contributions

P.J.E. designed the research and performed the experiments, analysed the data, wrote the manuscript. B.H.N. performed the experiments, analysed the data and provided intellectual input. J.C. and A.L.F. performed the experiments. W.Y.W. analysed the data. P.Y.G. and S.R.H. analysed data and provided intellectual input. J.D.O. designed the research, analysed the data and wrote the manuscript.

## Additional information

Correspondence and requests for materials should be addressed to J.D.O.

## Acknowledgments

We thank the donors for their blood donations. Authors would like to thank Anita Cummins, Kathleen Elford, Susan Morton and Ai Li Yeo for phlebotomy and Seiya Fukada for technical assistance.

## Supplementary Information

### Supplementary figures and tables

**Table S1.**
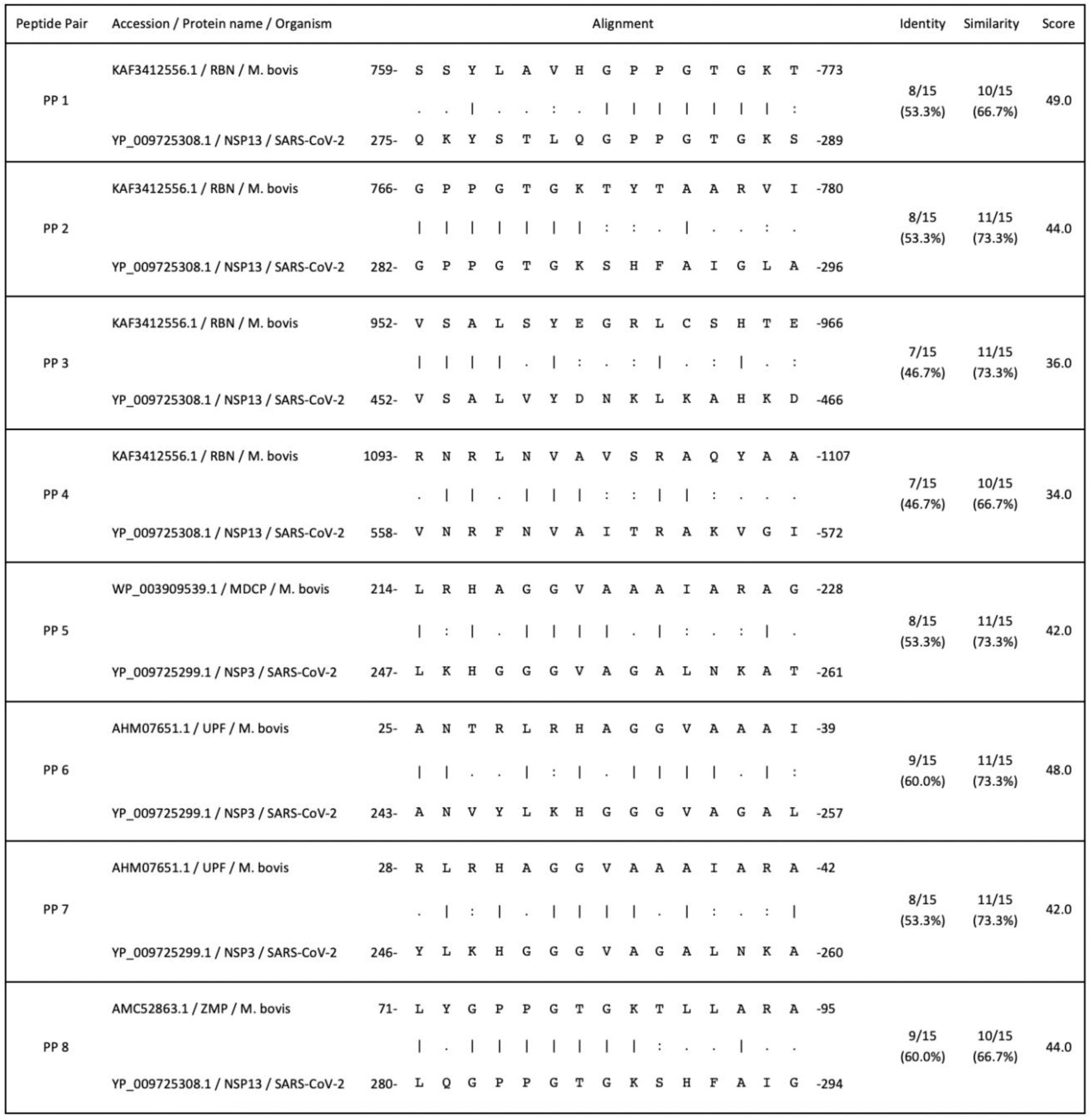
Sequence homology between BCG and SARS-CoV-2. Amino acid sequence alignment of the peptide pairs (PP) of BCG and SARS-CoV-2 used in this study including NCBI accession number, similarity, identity and BLOSUM62 matrix score. | - an identical amino acid match, : - a similar amino acid match,. – no match. RBN - RecB nuclease, MDCP - macro domain containing protein, UPF - UPF0189 protein, ZMP - zinc metalloprotease FtsH.

**Table S2a.** Donor characteristics.

|  |  |
| --- | --- |
| <u>Number:</u> | 20 |
| <u>Age:</u> | 21-42 |
| <u>Sex:</u> |  |
|  | Male: 50% (10/20) |
|  | Female: 50% (10/20) |
| <u>HLA typing:</u> |  |
|  | High resolution HLA-typed: 35% (7/20) |
|  | HLA-DR typed: 15% (3/20) |
|  | Not HLA-typed: 50% (10/20) |
| <u>BCG and TB status:</u> |  |
|  | BCG vaccinated: 50% (10/20) |
|  | BCG un-vaccinated: 50% (10/20) |
|  | Previous TB infection: 0% (0/20) |
| <u>Sars-CoV-2 status:</u> |  |
|  | IgG positivity: 0% (0/20) |
|  | IgM positivity: 0% (0/20) |

**Table S2b.**
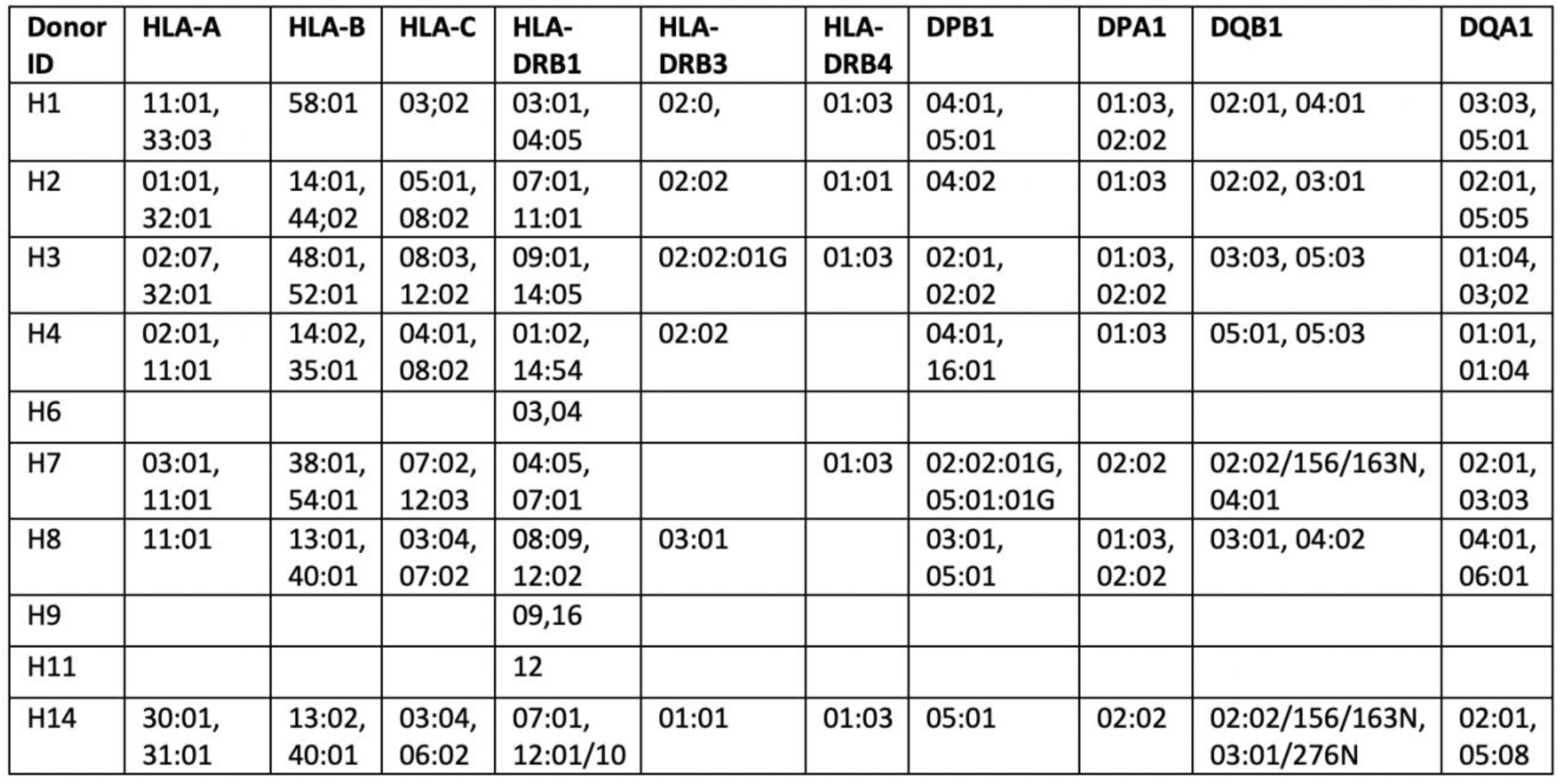
Donor HLA alleles used in this study.

| Donor ID | HLA-A | HLA-B | HLA-C | HLA-DRB1 | HLA-DRB3 | HLA-DRB4 | DPB1 | DPA1 | DQB1 | DQA1 |
| --- | --- | --- | --- | --- | --- | --- | --- | --- | --- | --- |
| H1 | 11:01, 33:03 | 58:01 | 03:02 | 03:01, 04:05 | 02:0, | 01:03 | 04:01, 05:01 | 01:03, 02:02 | 02:01, 04:01 | 03:03, 05:01 |
| H2 | 01:01, 32:01 | 14:01, 44:02 | 05:01, 08:02 | 07:01, 11:01 | 02:02 | 01:01 | 04:02 | 01:03 | 02:02, 03:01 | 02:01, 05:05 |
| H3 | 02:07, 32:01 | 48:01, 52:01 | 08:03, 12:02 | 09:01, 14:05 | 02:02:01G | 01:03 | 02:01, 02:02 | 01:03, 02:02 | 03:03, 05:03 | 01:04, 03:02 |
| H4 | 02:01, 11:01 | 14:02, 35:01 | 04:01, 08:02 | 01:02, 14:54 | 02:02 |  | 04:01, 16:01 | 01:03 | 05:01, 05:03 | 01:01, 01:04 |
| H6 |  |  |  | 03,04 |  |  |  |  |  |  |
| H7 | 03:01, 11:01 | 38:01, 54:01 | 07:02, 12:03 | 04:05, 07:01 |  | 01:03 | 02:02:01G, 05:01:01G | 02:02 | 02:02/156/163N, 04:01 | 02:01, 03:03 |
| H8 | 11:01 | 13:01, 40:01 | 03:04, 07:02 | 08:09, 12:02 | 03:01 |  | 03:01, 05:01 | 01:03, 02:02 | 03:01, 04:02 | 04:01, 06:01 |
| H9 |  |  |  | 09,16 |  |  |  |  |  |  |
| H11 |  |  |  | 12 |  |  |  |  |  |  |
| H14 | 30:01, 31:01 | 13:02, 40:01 | 03:04, 06:02 | 07:01, 12:01/10 | 01:01 | 01:03 | 05:01 | 02:02 | 02:02/156/163N, 03:01/276N | 02:01, 05:08 |

**Figure S1.**
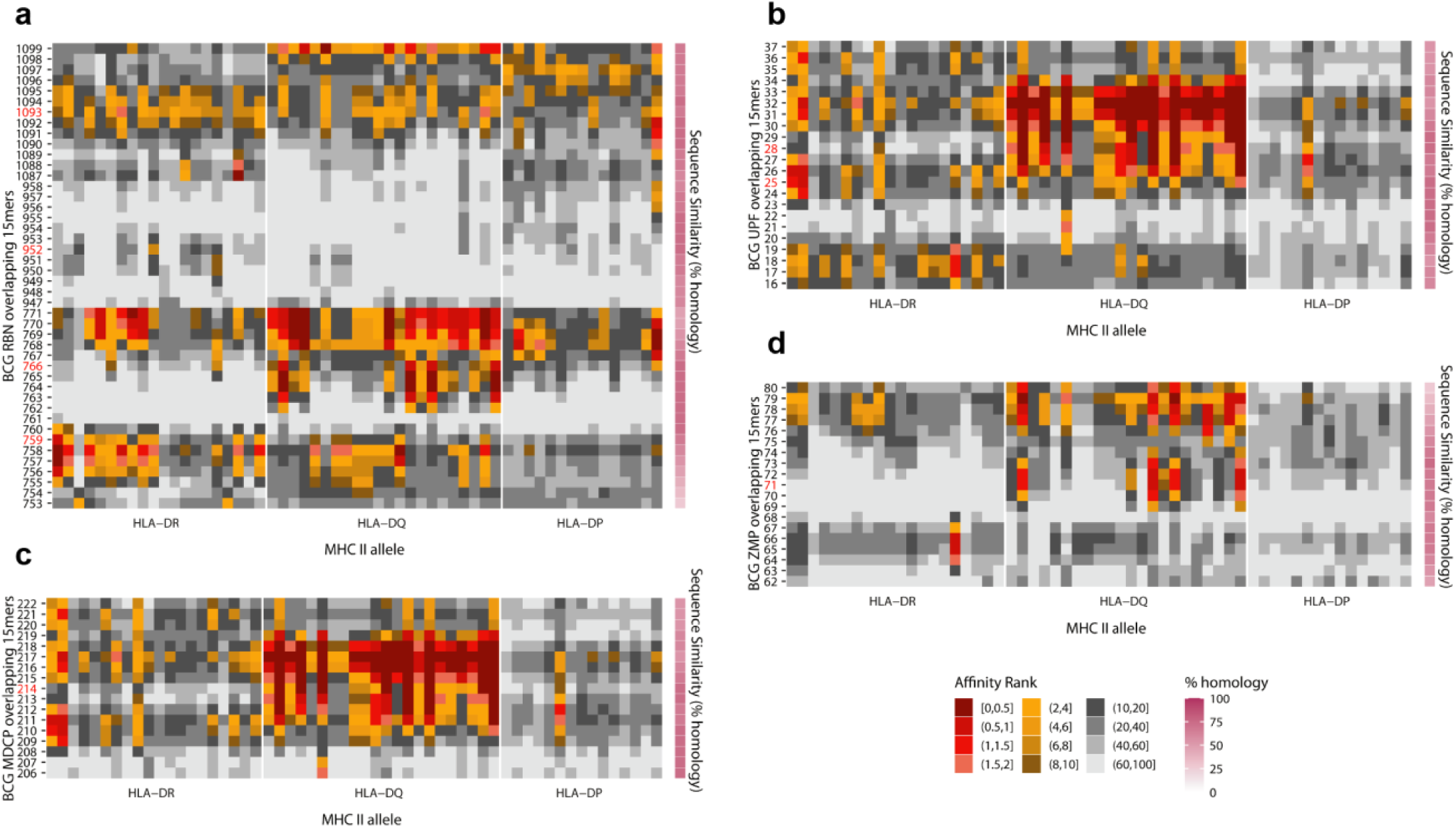
Regions of BCG-SARS-CoV-2 homology exhibit broad HLAII binding. Affinity rank score of BCG 15mers overlapping by 1 amino acid across the region of shared homology between BCG and SARS-CoV-2. Hotspots of high affinity overlap broadly with high regions of homology. Red gradient; strong peptide-MHC binder (affinity rank ≤ 2). Yellow gradient; peptide-MHC binder (affinity rank > 2 and ≤ 10). Grey gradient; non-binder (affinity rank < 10). Y-axis; number indicates the amino acid sequence start number of the respective 15mer. Red number indicates the 15mers analysed in this study. X axis; MHC class II alleles grouped into HLA-DR (n=20), HLA-DQ (n=22) and HLA-DP (n=15) isotype. The selected alleles are globally representative and include all alleles from HLA-typed donors used in this study. Pink gradient; Pairwise percent sequence similarity between BCG and SARS-CoV-2 15mers. a) BCG RecB nuclease (RBN) affinity rank binding scores. b) BCG UPF0189 protein (UPF) affinity rank binding scores. c) BCG macro domain containing protein (MDCP) affinity rank binding scores. d) BCG zinc metalloprotease FtsH (ZMP) affinity rank binding scores.

**Figure S2.**
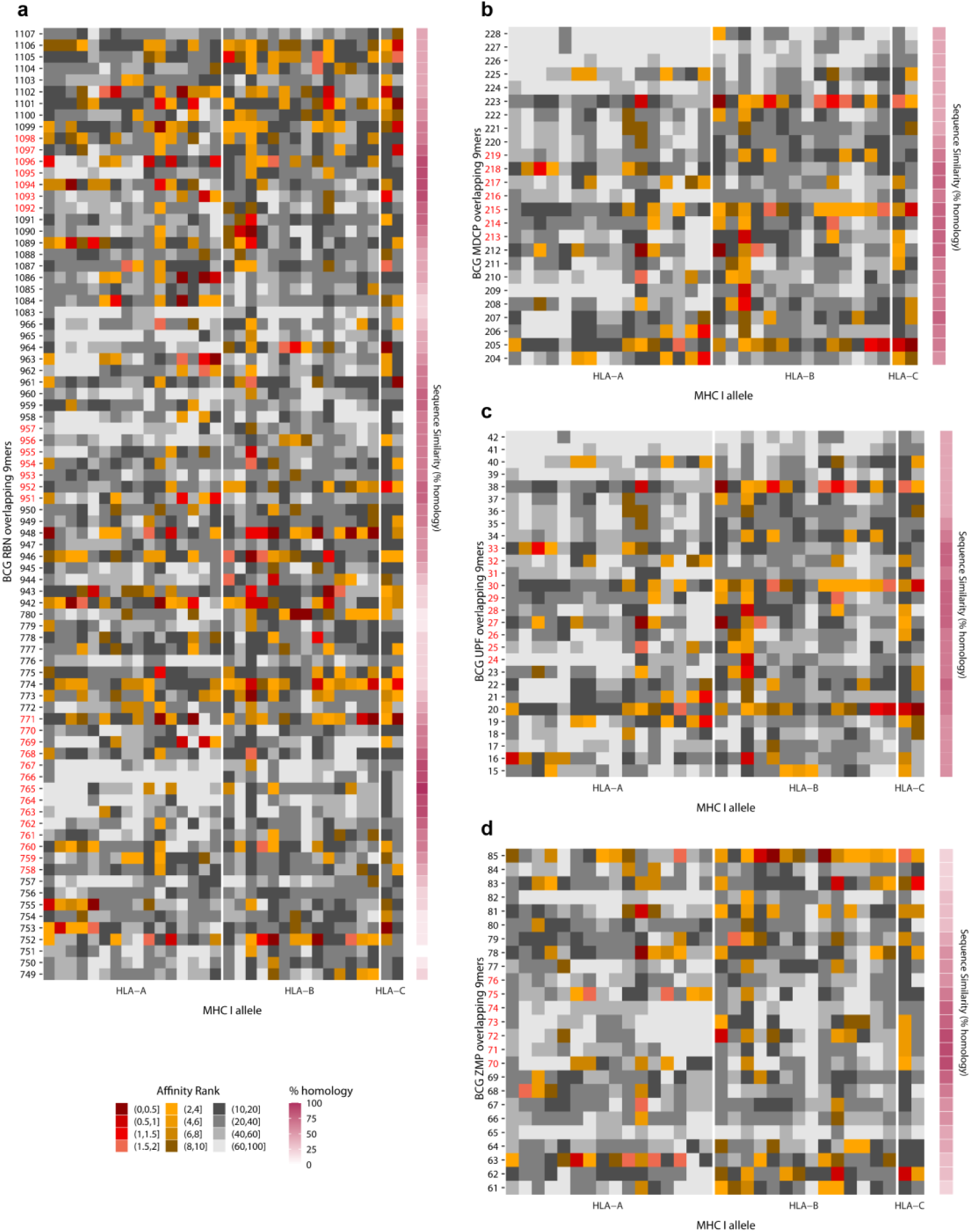
Regions of BCG-SARS-CoV-2 homology exhibit broad HLAI binding. Affinity rank score of BCG 9mers overlapping by 1 amino acid across the region of shared homology between BCG and SARS-CoV-2. Red gradient; strong peptide-MHC binder (affinity rank ≤ 2). Yellow gradient; peptide-MHC binder (affinity rank > 2 and ≤ 10). Grey gradient; non-binder (affinity rank < 10). Y-axis; number indicates the amino acid sequence start number of the respective 9mer. Red number indicates the overlapping 9mers contained within the 15mers analysed in this study. X-axis; MHC class I alleles grouped into HLA-A (n=16), HLA-B (n=14) and HLA-C (n=2) isotype. These alleles selected are globally representative and include the alleles from HLA-typed donors used in this study. Pink gradient; Pairwise percent sequence similarity between BCG and SARS-CoV-2 9mers. a) BCG RecB nuclease (RBN) affinity rank binding scores. b) BCG macro domain containing protein (MDCP) affinity rank binding scores. c) BCG UPF0189 protein (UPF) affinity rank binding scores. d) BCG zinc metalloprotease FtsH (ZMP) affinity rank binding scores.

**Figure S3:**
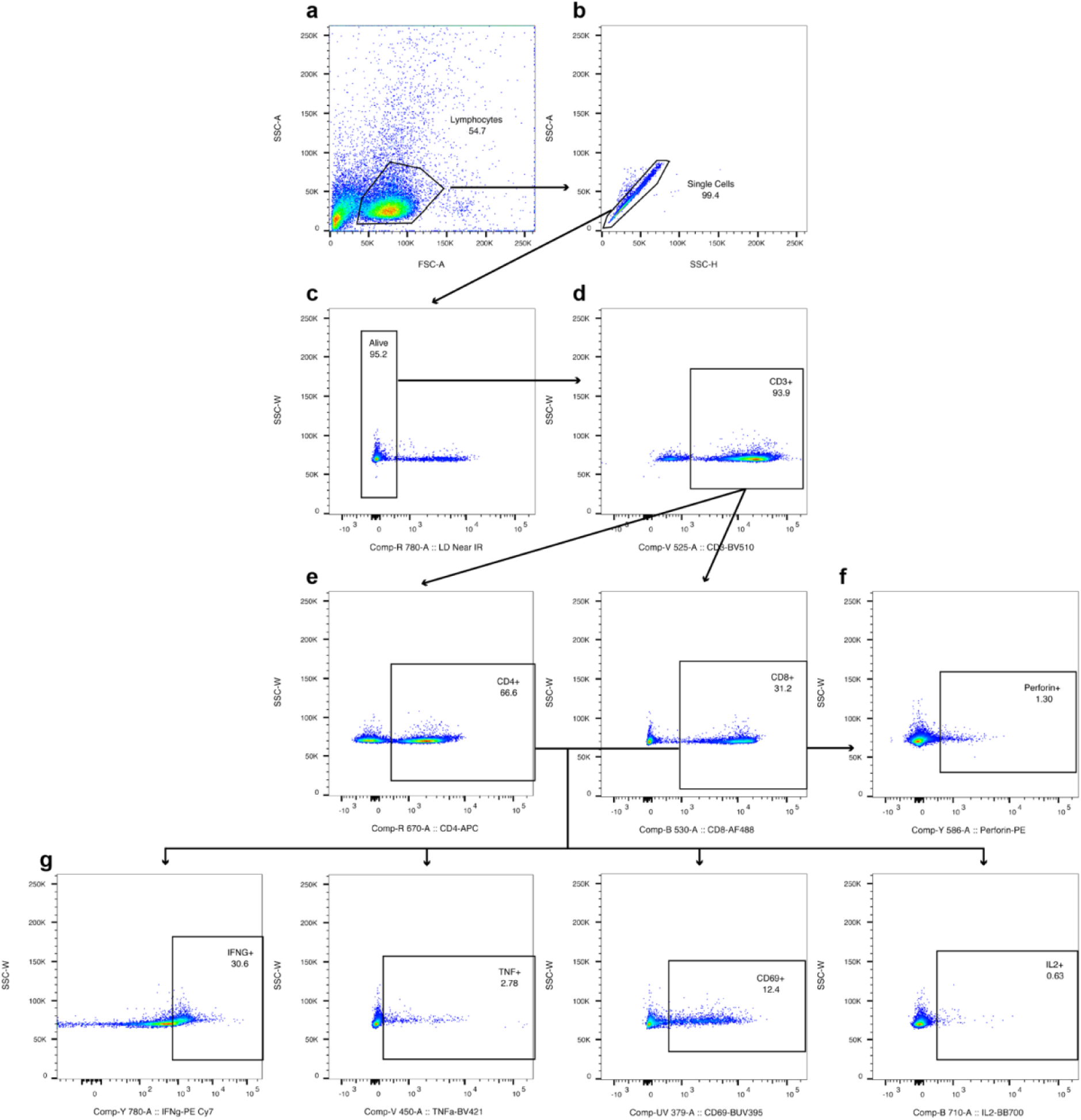
Flow Cytometry Gating Strategy for intracellular cytokine staining. Intracellular Cytokine Staining (ICS) panel. a) Forward scatter area (FSC-A) versus side scatter area (SSC-A) density plot gating the lymphocyte population. b) Side scatter height (SSC-H) versus SSC-A density plot gating the single cell population. c) Live/dead discrimination dye (LD Near IR) versus side scatter width (SSC-W) density plot gating the alive cell population. d) CD3 versus SSC-W density plot gating the CD3+ T cells. e) CD3+ T cells are separated into CD4+ and CD8+. f) Perforin positive gating is generated from the CD8+ parent population. g) IGNG, TNF, CD69 and IL-2 positive gates are generated from the CD4+ and CD8+ parent population. All fluorescence based gating is determined based on fluorescence minus one (FMO) controls.

**Figure S4:**
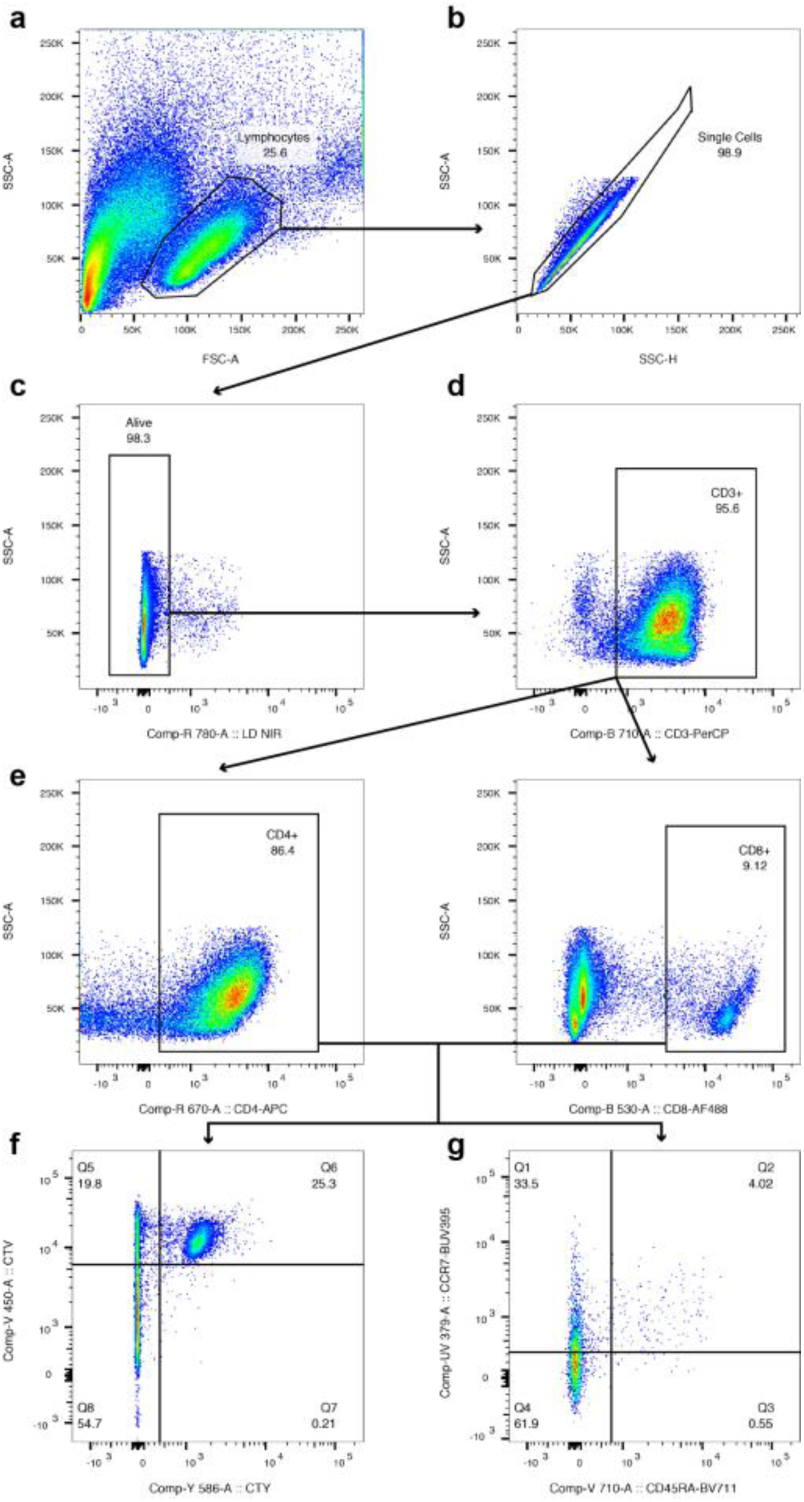
Flow Cytometry Gating Strategy for T cell proliferation and memory. T cell proliferation and memory gating strategy. a) Forward scatter area (FSC-A) versus side scatter area (SSC-A) density plot gating the lymphocyte population. b) Side scatter height (SSC-H) versus SSC-A density plot gating the single cell population. c) Live dead discrimination dye (LD Near IR) versus SSC-A density plot gating the alive cell population. d) CD3 versus SSC-A density plot gating the CD3+ T cells. e) The CD3^+^ cells are separated into CD4^+^ and CD8+ T cells. f) The proliferation dyes Cell Trace Yellow (CTY) versus Cell Trace Violet (CTV) density plots are quadrant gated based on proliferation after priming and restimulation. Q5 – CTY^low^ CTV^high^ T cells that proliferated after priming but not after restimulation. Q6 – CTY^high^ CTV^high^ T cells that did not proliferate after priming or restimulation. Q7 CTY^high^ CTV^low^ T cells that did not proliferate after priming but proliferated after restimulation. Q8 – CTY^low^ CTV^low^ T cells that proliferated both after priming and restimulation. g) The CD45RA versus CCR7 density plots of the CD4^+^or CD8^+^ parent populations are quadrant gated separating the Q1 - CD45RA^-^ CCR7^+^ (T central memory cells), Q2 - CD45RA^+^ CCR7^+^ (Naïve T cells), Q3 - CD45RA^+^ CCR7^-^ (TEMRA cells), and Q4 - CD45RA^-^ CCR7^-^ (T effector memory cells). All fluorescence based gating except CTY and CTV is determined based on fluorescence minus one (FMO) controls. CTV and CTY gating is based on the point at which the first cell division took place visible by fluorescence dye dilution.

**Figure S5.**
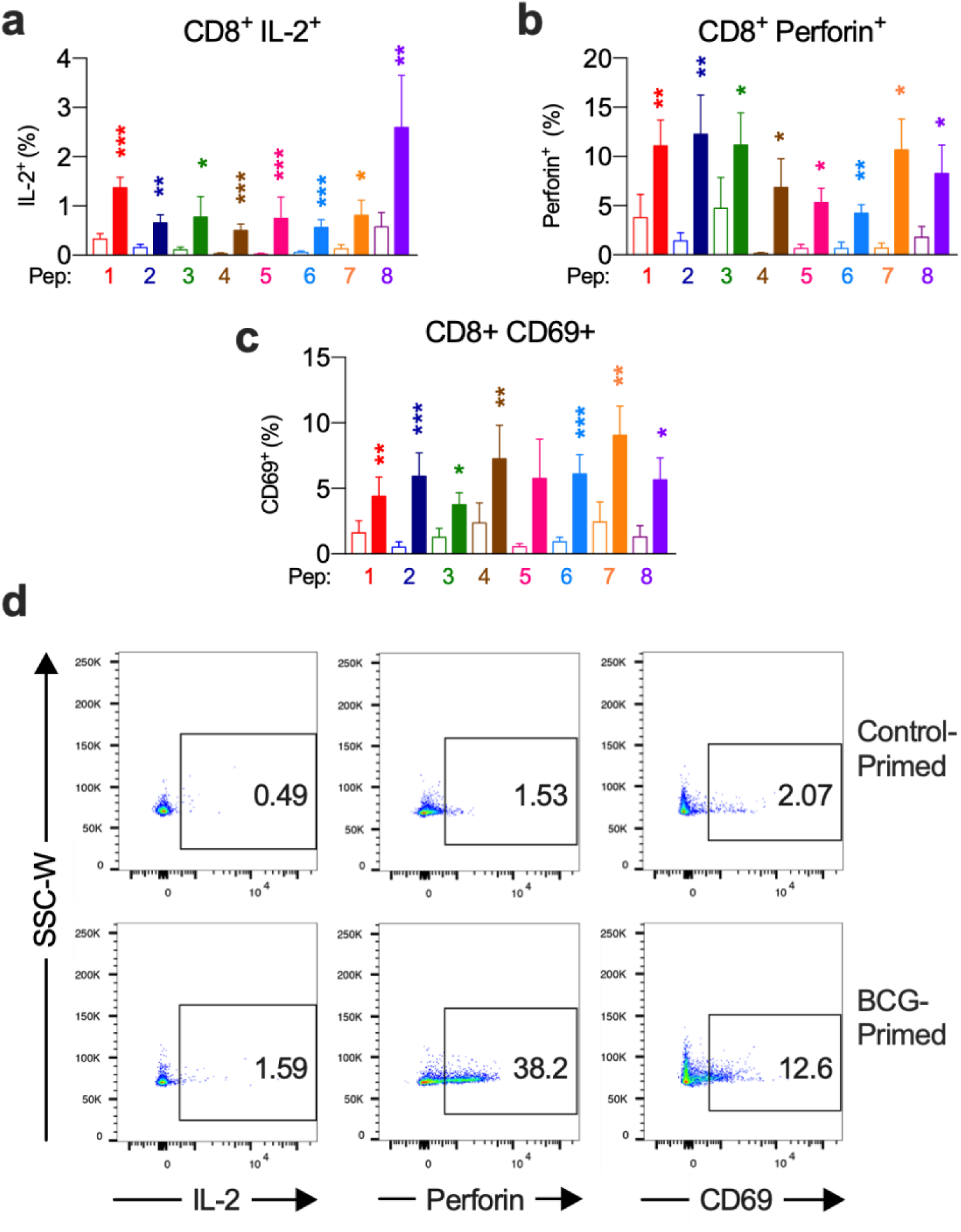
BCG priming enhances IL-2, Perforin and CD69 CD8 T cell responses against SARS-CoV-2. BCG-peptide 1-8 primed (shaded bars) or control primed using irrelevant peptide CLIP_103-117_ (unshaded bars) CD3^+^ T cells were restimulated with SARS-CoV-2-peptide-homologue-pulsed dendritic cells for 6hr and analysed by intracellular cytokine staining. Responders were selected as per methods section for analysis. *P < 0.05, **P < 0.01, ***P < 0.001 by Wilcoxon matched-pairs signed rank test. a) CD8^+^ IL-2^+^ responses (n=7-13). b) CD8^+^ Perforin^+^ responses (n=6-14). c) CD8^+^ CD69^+^ responses (n=5-12). d) Representative IL-2, perforin and CD69 dot plots of a responder control-primed (top) and BCG-primed (bottom) donor.

**Figure S6.**
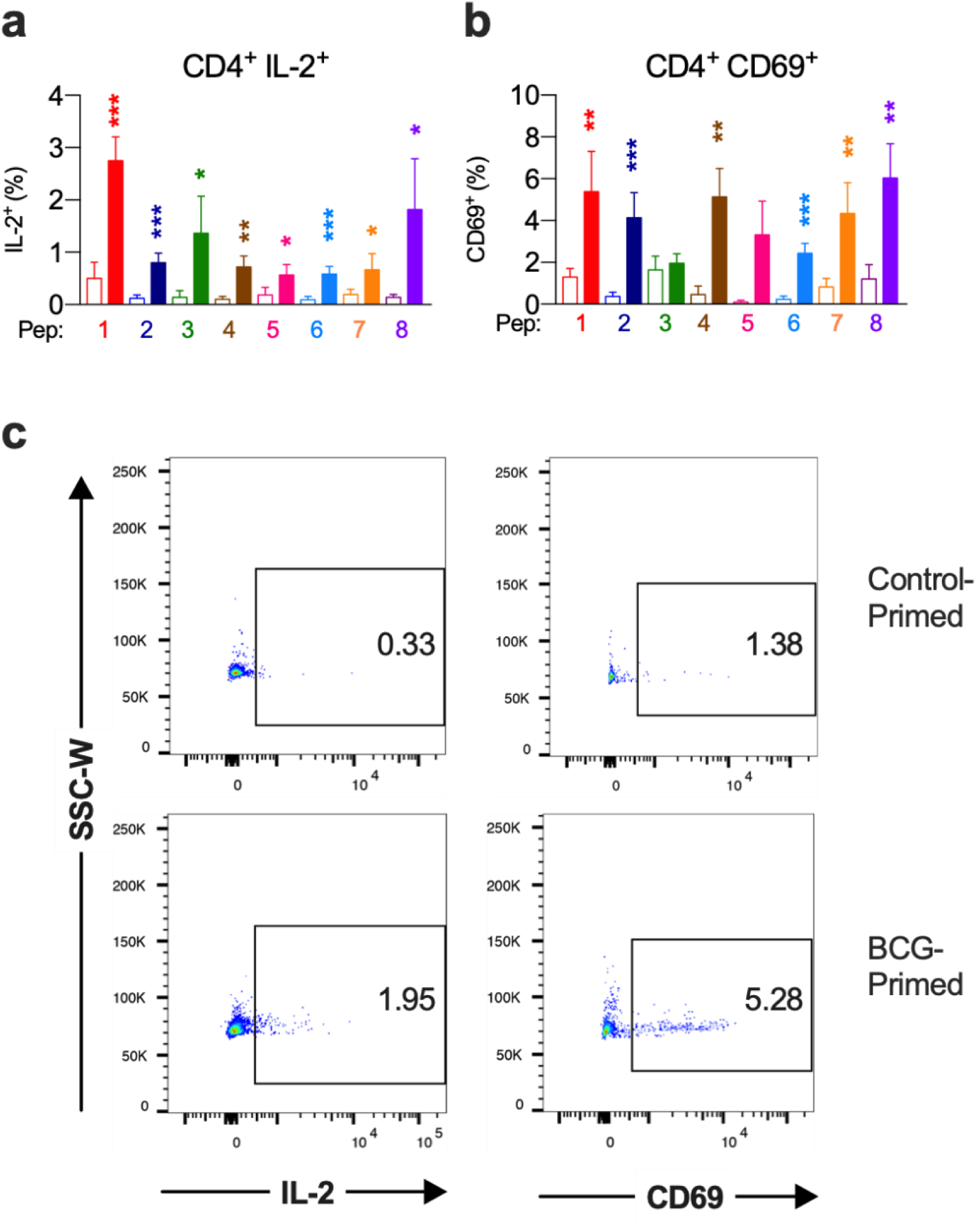
BCG priming enhances IL-2 and CD69 CD4 T cell responses against SARS-CoV-2. BCG-peptide 1-8 primed (shaded bars) or control primed using irrelevant peptide CLIP_103-117_ (unshaded bars) CD3^+^ T cells were restimulated with SARS-CoV-2-peptide-homologue-pulsed dendritic cells for 6hr and analysed by intracellular cytokine staining. Responders were selected as per methods section. *P < 0.05, **P < 0.01, ***P < 0.001 by Wilcoxon matched-pairs signed rank test. a) CD4^+^ IL-2^+^ responses (n=7-13). b) CD4^+^ CD69^+^ responses (n=8-13). c) Representative IL-2 and CD69 dot plots of a responder control-primed (top) and BCG-primed (bottom) donor.

**Figure S7.**
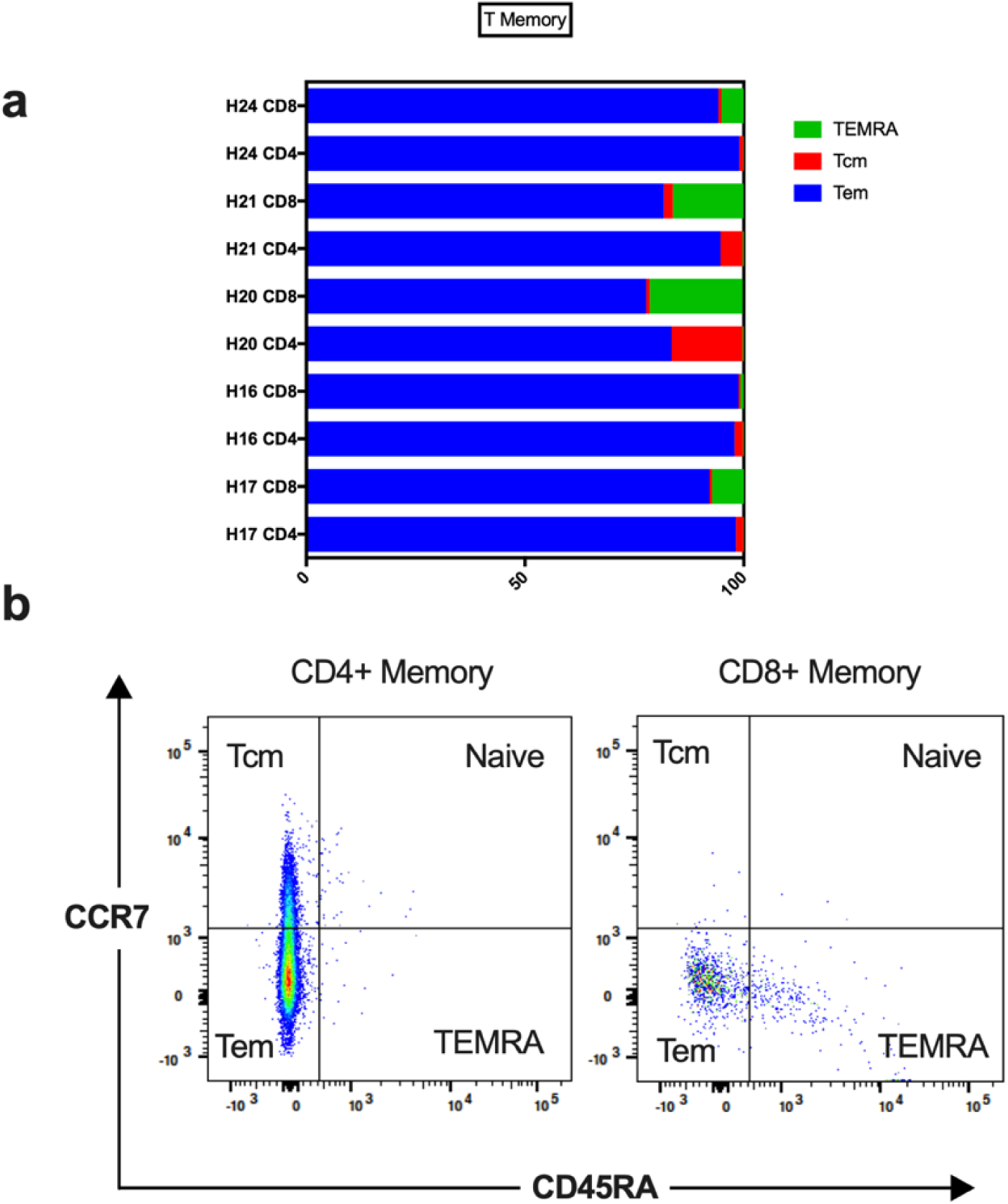
BCG-stimulated T cells express memory phenotypes. BCG-peptide-stimulation of T cells produces > 99% T memory phenotype after 16 days co-culture. T effector memory (Tem) is the dominant memory phenotype for both CD4^+^ and CD8^+^ T cells. CD8^+^ T cells exhibit a subpopulation of T effector memory re-expressing CD45RA (TEMRA) with minimal T central memory (Tcm). CD4^+^ T cells exhibit a subpopulation of Tcm with minimal TEMRA. a) Composition of CD4^+^ and CD8^+^ T cell memory subpopulations in BCG UPF0189 protein_25-39_ (from peptide pair 6) primed T cells, cultured for 16 days; 7 days with peptide and 9 days without peptide. A representative sample of 5 individuals (from n = 20) from 1 peptide pair (from n=8). b) Representative dot plots of CD45RA versus CCR7 expression of the proliferated CD4^+^ or CD8^+^ T cells based on Cell Trace Violet dilution. T memory phenotype was characterised as three subpopulations by expression of CD45RA and CCR7. Tem of phenotype CD45RA^-^ and CCR7^-^. Tcm of phenotype CD45RA^-^ and CCR7^+^. TEMRA of phenotype CD45RA^+^ CCR7^-^. Non-memory T naïve of phenotype CD45RA^+^ CCR7^+^.

**Figure S8.**
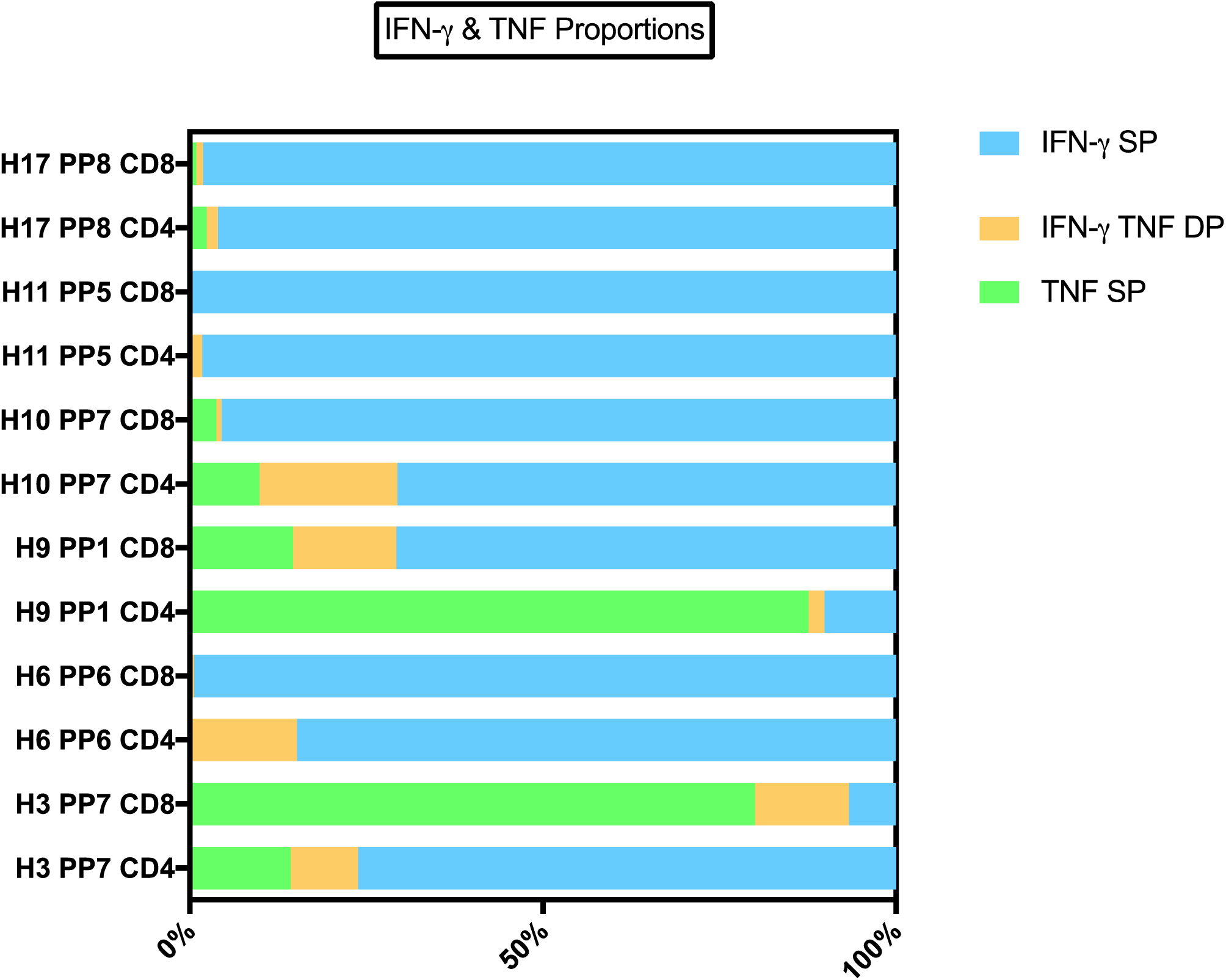
TNF IFN-γ proportions of responder individuals. BCG-primed individuals who responded to SARS-CoV-2-peptide restimulation mostly exhibited a strong IFN-γ signature with a lower proportion of TNF, although some individuals showed a dominant TNF response. Across individuals and peptide pairs, a variable proportion of IFN-γ SP, TNF SP and IFN-γ, TNF DP was observed. A selection of 6 individuals CD4+ and CD8+ responses (of N = 20 tested) against 5 peptide pairs (PP) of 8 peptide pairs tested. IFN-γ single positive (SP) – proportion of cells producing IFN-γ and not TNF. TNF SP – proportion of cells producing TNF and not IFN-γ. IFN-γ TNF double positive (DP) – proportion of cells producing TNF and not IFN-γ.

## Notes

### Competing Interest Statement

Dr. Ooi reports grants from Monash Health Foundation,  during the conduct of the study; no financial relationships with any organizations that might have an interest in the submitted work in the previous three years; no other relationships or activities that could appear to have influenced the submitted work.

### Funding Statement

Dr. Ooi reports grants from Monash Health Foundation during the conduct of the study. No other payments or services from a third party were received at any time for any aspect of the submitted work.

### Author Declarations

Monash University Human Research Ethics Committee project ID 25834.

